# Quantitative Reality: A Physical Ground Truth to Assess Heterogeneity Beyond Visual Limits in Molecular Imaging

**DOI:** 10.1101/2025.11.26.25341036

**Authors:** Sai Kiran Kumar Nalla, Quentin Maronnier, John A. Kennedy, Olivier Caselles

## Abstract

**Background:** Accurate quantification of intratumoral heterogeneity is essential for validating radiomics features and AI models in oncology. However, standard physical phantoms typically utilize homogeneous compartments, lacking the spatially complex uptake patterns required to rigorously benchmark these metrics against a known reality. To address this, we introduce porous 3D-printed inserts for simulating controlled spatial heterogeneity.

**Methods:** Modular grid inserts (17, 22, 28 mm targets) were developed with contrast of 4:1. Nine heterogeneity models, simulating progressions from uniform uptake to multifocal to necrosis, were imaged on a 32cm Omni Legend PET/CT system. Two different tracers: [^18^F]FDG ([^18^F]-2-fluoro-2-deoxy-d-glucose) and [^68^Ga] (68-Gallium) were used on different days. Quantitative performance was assessed using conventional Standardized Uptake Values (SUV) and Target-to-Background Ratios (TBR), alongside the Gini Index (GI) to quantify spatial heterogeneity and radiotracer distribution inequality.

**Results:** The phantom demonstrated high consistency, with reference solution variability remaining under 2.2% across layers. In 28 mm inserts, experimental TBR_max_ values fell within 10% of the theoretical 4:1 design ratio. GI successfully quantified structural complexity, ranging from 0.05 in homogeneous regions to 0.30 in necrotic targets. Cross-tracer analysis demonstrated robust reproducibility with a mean bias of <10%. While Partial Volume Effects reduced visual distinctness in 17 mm geometries, quantitative metrics successfully preserved the intended pathological trends.

**Conclusion:** This study establishes a foundational ground truth for heterogeneous uptake in PET. By providing defined physical standards validated by quantitative metrics, it enables the robust validation of segmentation algorithms and radiomics features even when spatial resolution limits visual assessment.

## 1. Introduction

Radiotracer uptake is a defining characteristic of lesions in nuclear medicine playing a crucial role in quantification, feature extraction, and characterization **[1–3]**. In clinical practice, however, lesions rarely demonstrate uniform uptake; instead, they often display heterogeneous patterns such as necrotic cores, peripheral gradients, or multifocal regions **[3–5]**. Quantitative metrics derived from such patterns, including radiomic features, form the basis for advanced Artificial Intelligence (AI) models used in prognostication and personalized medicine. Crucially, in some tumours underlying heterogeneity can be present as low-contrast or visually indistinct patterns, leading to the risk of underestimating the true biological complexity. Hence, accurate quantification of this heterogeneity is essential for treatment response evaluation and multi-centre harmonization **[6–8]**. Physical phantoms that are widely used for performance testing and standardization of imaging **[9,10]** typically represent homogeneous activity distributions. Despite clinical importance, there remains a lack of practical tools for studying spatial heterogeneous uptake patterns under controlled and repeatable conditions.

In this context, Three dimensional (3D) printing offers a flexible platform for phantom development. Precise control of geometry and material composition were previously achieved for nuclear medicine and radiation therapy based studies **[11–13]**. Existing 3D printed phantom designs range from organ models derived from patient segmentations **[14–16]** to emulated lesions with controllable uniform uptake characteristics **[15,17–21]**. From the perspective of spatial heterogeneity, compartment-based phantoms have been created based on regions with different activity levels **[18,22]**, but they require labour-intensive preparation, careful manual filling, and risk leakage or cross-contamination. Approaches that incorporate radioactivity directly into the printing resin have been explored as an alternative but only for uniform targets **[23,24]**. Among existing strategies, permeable grid-based designs are particularly promising **[15,17,19]**. These grids alternate between regions smaller than the scanner’s spatial resolution, producing an averaged effect on the reconstructed image. This principle allows for fine control over tracer distribution using single injection without the need for complex compartmentalization or multiple manual filling steps. Such inserts have been proven to be conceptually effective with strong transferability and reproducibility across facilities **[25,26]**. However similar to direct radioactive printing, they were tested primarily to simulate uniform targets or organ-level phantoms. There is therefore a clear need for a simple, reproducible, and scalable approach to generate heterogeneous activity distributions while maintaining quantitative interpretability and potential for cross-site standardization.

In this work, we present a grid-based 3D-printed phantom inserts with controlled spatial heterogeneous uptake characteristics while preserving protocol simplicity and quantitative robustness. The proposed design incorporates homogeneous regions, “no-target” zones (background or non-avid tissue), necrotic areas (near zero uptake) and multifocal sub-regions representing varied activity. To assess consistency, a single phantom was imaged using two different radiotracers on the same PET/CT (Positron Emission Tomography / Computed Tomography) system. The dual-tracer framework is geared to evaluate design robustness rather than quantitative cross-tracer performance analysis. With the help of conventional PET metrics, we further examine whether the design yields quantifiable heterogeneity, even in scenarios where such variation is not visually apparent.

## 2. Methods

### 2.1 Base Phantom Design

The basic design of phantom inserts is adapted from **[19]** in which all experiments were conducted within a standard Jaszczak cylinder phantom (total fill volume ≈ 5.5 L for this study). Each insert is based on a 40 mm porous cube in which spherical voids (17, 22, and 28 mm) were subtracted using Autodesk Fusion 360 (Autodesk Inc, San Francisco, CA, USA). These penetrable voids act as targets **(Fig.1a)**, allowing to be filled with the tracer solution and reach intended activity concentration (AC) following **Eq. 1, 3**. The **solidity** *S* (or infill percentage) of the background mesh **(Fig.1a)** represents the fraction of solid material and therefore controls tracer dilution. For a given background solidity (S_bkg_), the effective tracer activity concentration within the mesh is given by **Eq.2**. A 3 × 3 array of the inserts as illustrated in **Fig.2** were placed concentrically within the cylinder phantom leading to 3 distinct regions as shown in **Fig 1**.

**Fig.1.**
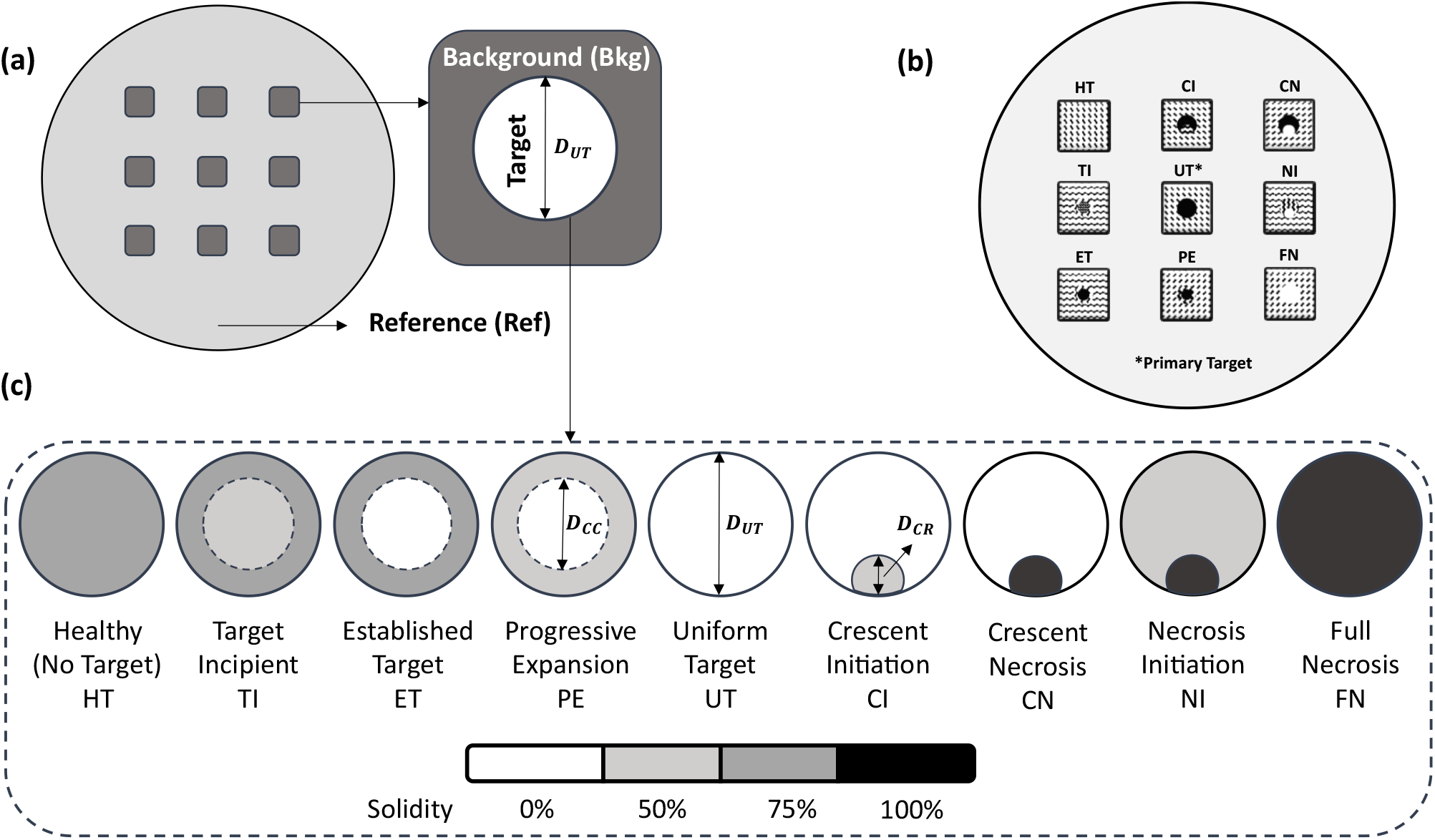
**(a)** Different regions of the custom phantom – Reference refers to the radiotracer + water solution; **(b)** 3 x 3 Layout used for each target size layer; **(c)** 9 targets types designed to simulate lesions with multi-uptake patterns, Solidity used to create dilution for different regions are represented using a colour bar (3 base controllable design parameters - D_UT_ : Diameter of uniform target, D_CC_ : Diameter of central core, D_CR_ : Diameter of crescent region. Design configurations are also detailed in appendix (**Table A1**).

**Fig.2.**
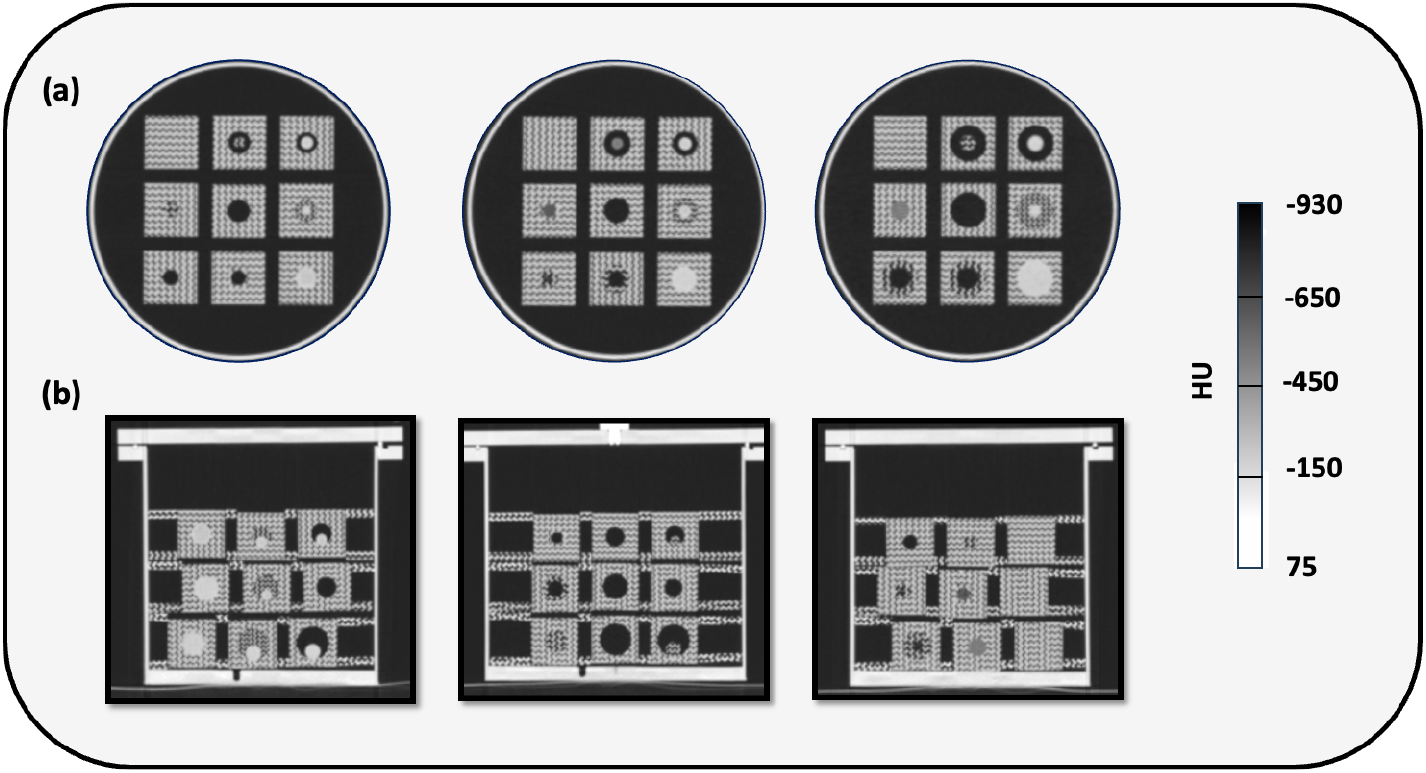
**(a)** Central Axial Slices for CT Air for 17mm (Left), 22mm (Middle), 28mm (Right); **(b)** Coronal view illustrating the crescent shapes of different infills and sizes

- Reference (Tracer uptake in the water outside of the inserts filled inside the phantom, used as reference to benchmark target uptake)

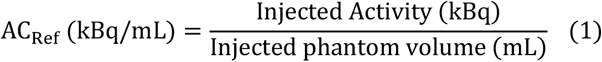
- Background mesh (Representing dimmer tissue emulation)

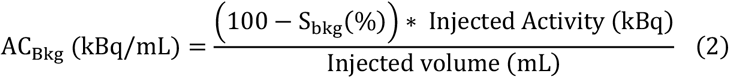
- Targets (Uptake in the water within the voids - Representing hot lesion emulation)

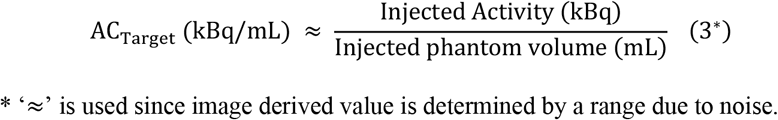

Target to background ratio (TBR) of these inserts with targets is defined using below **Eq.4**. Under ideal tracer accumulation within the void, AC_ref_ ≈ AC_target_ leading to AC_Bkg_ ≈ TBR x AC_ref_ ∼ TBR x AC_target_. For this study, a 75% infill (solidity) was used for the background mesh. This choice was made to achieve a higher theoretical TBR_insert_ ∼ 4, pushing beyond the limits of previous grid-based studies **[15,17,26]**, while ensuring sufficient range for creating heterogeneous patterns.

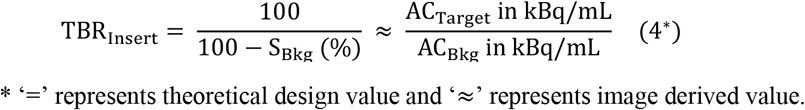

### 2.2 Heterogeneous Target Design

Spatial heterogeneity simulation is achieved through introduction of a secondary sphere of different dimensions (**Appendix**) within these voids by another Boolean operation thus creating two nested regions (shown in **Fig. 1c**):

- The **primary region (region A)**, representing the highest tracer uptake
- The **secondary region (region B)**, representing reduced or necrotic activity

By varying the infill of these two regions (using either S: 0%, 50% or 100% representing TBR ∼ 1, 2, Infinity) different uptake patterns can be generated to emulate various stages of lesion evolution - from healthy tissue to full necrosis or multifocal uptake. Nine distinct models were created for each base insert configuration (i.e., each uniform void size), resulting in a total of 27 inserts. The spatial arrangement of for each layer within the phantom is shown in **Fig.1b**. It is to be noted that the TBR for this new heterogeneous target now changes to a voxel based spatial function defined using **Eq.5**.

The TBR_insert_ defined earlier is only used as a reference design parameter for cube identification and phantom positioning. Since background mesh and healthy insert share same infill percentage, the latter is always used in this study for quantitative calculations to have uniform assessment framework in case of delineation difficulties.

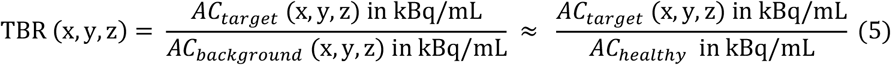

All designs were exported as .OBJ files and printed using a Prusa XL 3D printer using parameters detailed in **[19]** in acrylonitrile butadiene styrene (ABS). Printing success was verified via CT imaging (1.25 mm slice thickness).

### 2.3 PET/CT Imaging Parameters & Data Analysis

The phantom was imaged on a Discovery OMNI Legend 32cm PET/CT system (*GE Healthcare, Chicago, IL, USA*) using two different tracers, [^18^F]FDG ([^18^F]-2-fluoro-2-deoxy-d-glucose) and [^68^Ga] (68-Gallium), to evaluate design robustness and visualization consistency. The phantom was filled with soap & water solution followed FDG injection (approximately 6 hours post filling) for first acquisition. The phantom was then left undisturbed overnight for sufficient decay of the tracer. The following day Ga injection was carried out using same protocol for the second acquisition. The scan protocol was adjusted based on the tracer when necessary as shown in **Table 1** but following the recommendations from **[26]**. The attenuation CT (CTAC) parameters were maintained same as **[26]**. The acquired PET data was extracted using a semi-automatic workflow. Two distinct quantitative extractions were performed. First, for overall heterogeneity analysis, a standardized spherical Volume of Interest (VOI) with a diameter (+10% tolerance) than the insert’s nominal design size (17, 22, 28mm), was automatically placed at the centroid of each insert. Second, for quantitative assessment of the primary region, an iso-surface segmentation analysis was performed on the most active regions within the same reference VOIs. Segmentation statistics were extracted as a function of the iso-surface threshold value. The results from the 75% iso-surface threshold (Corresponding to highest S: 75%) are utilised for main analysis.

**Table. 1.** Acquisition & Reconstruction Parameters.

| Acquisition Parameter | $^{18}F$-FDG | $^{68}Ga$ |
| --- | --- | --- |
| Bed Positions | 1 |  |
| Matrix Dimensions | 384 x 384 |  |
| Field of View | 600 mm |  |
| Slice Thickness | 2.07mm |  |
| Pixel Dimensions | 1.56mm x 1.56mm |  |
| No. of Slices | 153 |  |
| Voxel Volume | 5.05mm<sup>3</sup> |  |
| Injected Activity | 68.38 MBq | 93.04 MBq |
| Injection to Scan relaxation time | 42min | 25min |
| Scan Duration | 3 x 300s (50MBq)<br>3 x 600s (30MBq) | 3 x 600s (50MBq)<br>3 x 600s (30MBq) |
| Reconstruction Algorithm | QCHD (BPL*) |  |
| Reconstruction $\beta$ parameter | 650 | 750 |
| *Bayesian penalized-likelihood reconstruction with regularization parameter beta |  |  |

*NOTE: This preprint reports new research that is not certified by peer review and should not be used to guide clinical practice.*

Five different quantitative PET metrics were calculated from the extracted data, namely

- Standardized uptake value (SUV) for different regions from VOI data is given by

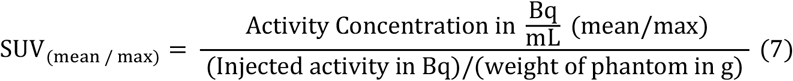
- Gini index (GI) was utilised as heterogeneity measure within the target volume, where a coefficient of 0 represents perfect homogeneity and 1 represents maximal heterogeneity **[18]**.

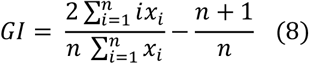

Where n is total No. of voxels in the VOI, x_i_ is voxel intensity of i-th voxel sorted in ascending order & i is the rank of the voxel in the sorted list
- Target to background ratio - TBR_quant_ for segmentation (i.e. Hottest region) given by

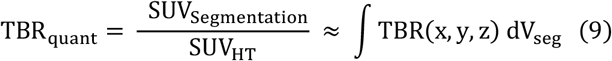

Statistical tests were performed to evaluate discrepancies. First, a Wilcoxon test was used to compare the extracted VOI data between each target type within each diameter group with a significance level of 0.05. The goal of Wilcoxon test is to quantify the combinations fail in similarity significance test (the higher the better). Second, a Bland-Altman analysis was conducted to assess the accuracy and potential bias. All data extraction and statistical analyses were conducted using MATLAB (*MathWorks INC, MA, USA*) and JASP (*JASP Team, Netherlands*).

## 3. Results

CT air performed to confirm layer positioning and print success is presented in **Fig 2**.

### 3.1 Visual and Quantitative Uptake Assessment

The 3D-printed inserts successfully generated heterogeneous uptake patterns on the reconstructed PET data for both tracers (**Fig 3 & 4**). The nine designed uptake patterns, including uniform targets (UT), necrotic cores (e.g., NI, FN), and crescent-shaped regions (CI, CN) were most clearly represented in the largest (28 mm) inserts for both tracers. The smaller 22 mm inserts showed some intended patterns, though they were less distinct, while for the 17 mm inserts, these heterogeneous patterns (such as crescents) were visually indiscernible due to significant partial volume effects. The visual consistency between the ^18^F-FDG and ^68^Ga acquisitions was reasonably high. Mean Hounsfield (HU_mean_) values within the VOIs on the CTAC varied between -3.2±6.4 to -38±20.4 for Fluorine and -3.1±4.6 to -31±13.4 for Gallium ensuring no presence of major air bubbles at scan resolution affecting the quantitation, falling within range prescribed by **[26]**. Uniform mixing of radiotracer was also confirmed based on layer wise variability as presented in **Table 2**.

**Table. 2.** SUV_mean_ for Reference Solution across layers.

| Layer No | $^{18}\text{F}$-FDG | | Maximum Variability in (%) | $^{68}\text{Ga}$ | | Maximum Variability in (%) |
| --- | --- | --- | --- | --- | --- | --- |
|  | 50 MBq | 30 MBq |  | 50 MBq | 30 MBq |  |
| 1 (17mm) | $0.94\pm 0.002$ | $0.92\pm 0.003$ | 2.12% for 50MBq<br>1% for 30MBq | $0.91\pm 0.003$ | $0.91\pm 0.001$ | 1% for 50MBq<br>2.15% for 30MBq |
| 2 (22 mm) | $0.92\pm 0.002$ | $0.92\pm 0.003$ | | $0.92\pm 0.003$ | $0.92\pm 0.002$ | |
| 3 (28 mm) | $0.93\pm 0.002$ | $0.91\pm 0.003$ | | $0.92\pm 0.003$ | $0.93\pm 0.003$ | |

*NOTE: This preprint reports new research that is not certified by peer and should not be used to guide clinical practice.*

**Fig.3.**
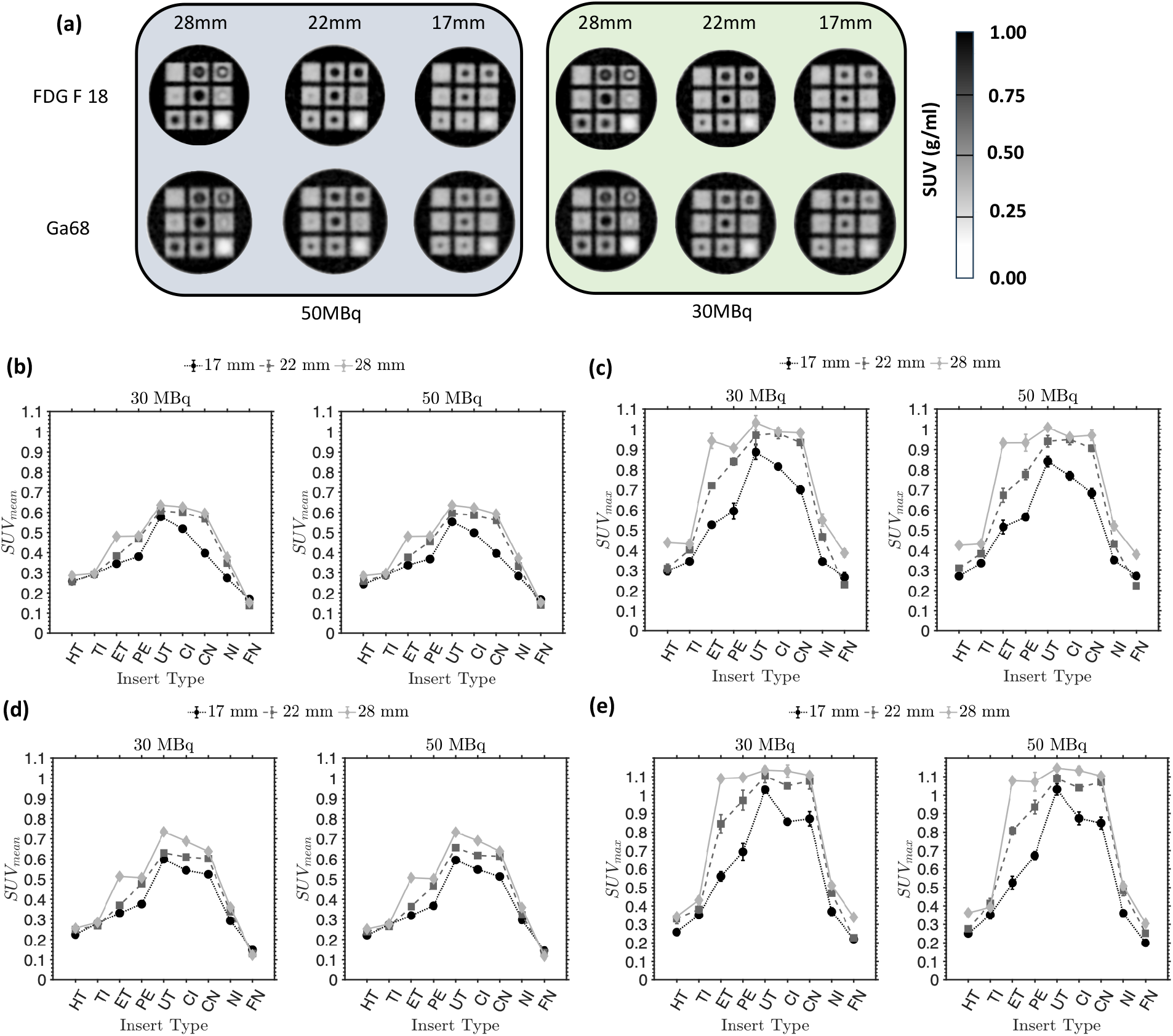
**(a)** PET axial slices of 3 layers for different activity concertation and tracers; Mean and Max SUV values as function of insert design type for (**b-c)** ^18^F-FDG and **(d-e)** ^68^Ga

**Fig.4.**
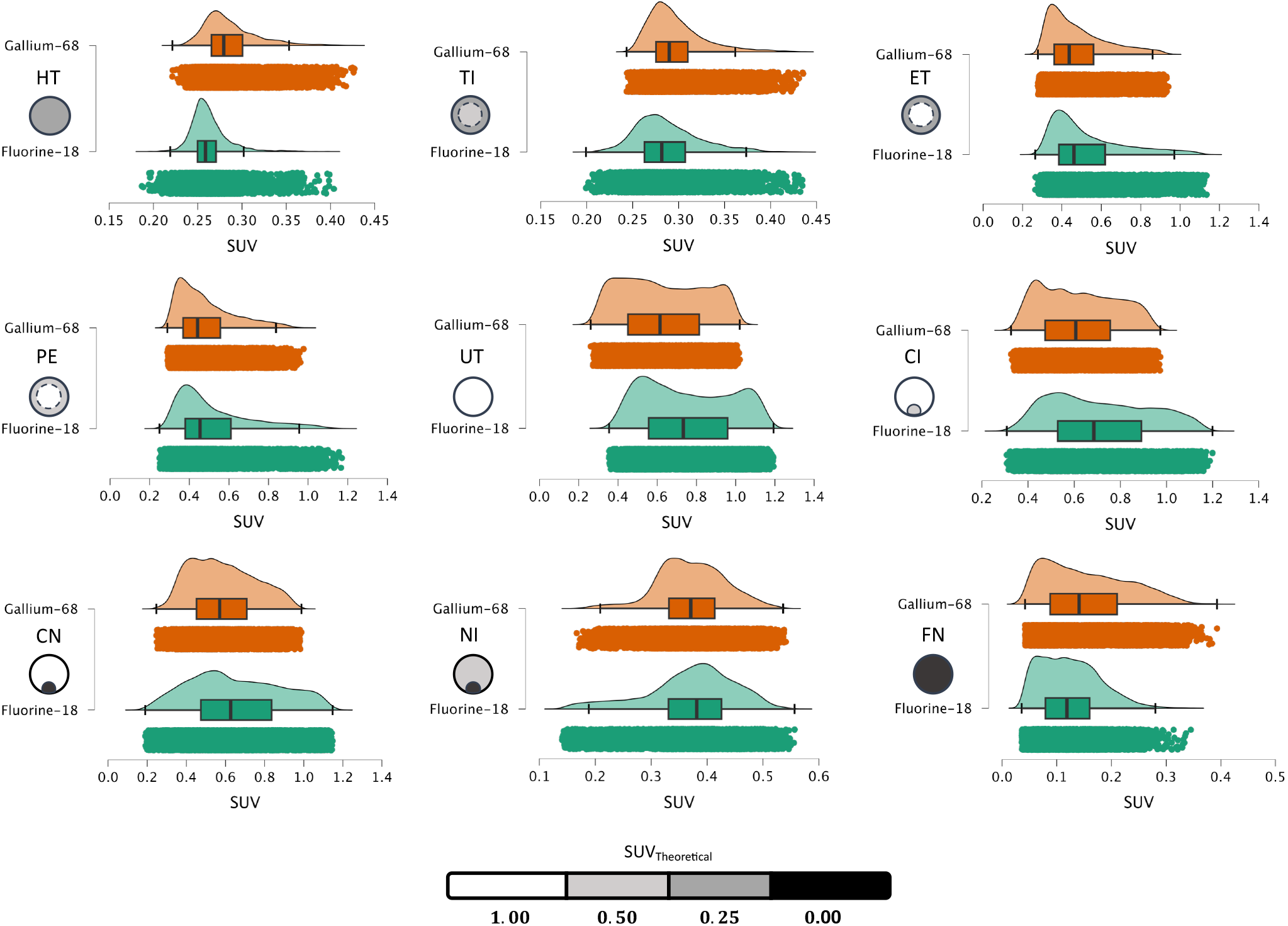
Kernel Density Estimation (KDE) histogram plots from VOI data for the 28 mm inserts from the 50 MBq acquisition. Each plot shows the distribution of voxel-level SUV values for both ^18^F-FDG (Orange), ^68^Ga (Green)

Quantitative analysis of SUV_mean_ and SUV_max_ for each insert type demonstrated adherence to the design principles (**Fig 3b-e**). The profiles followed the expected pathological progression: uptake was minimal in the ‘Healthy’ (HT) insert, increased through the ‘Incipient’ (TI), ‘Established’ (ET) and ‘Expansion’ (PE) stages, and peaked for the ‘Uniform Target’ (UT) and ‘Crescent Initiation’ (CI) inserts. As the simulated necrosis progressed (‘Crescent Necrosis’ (CN), ‘Necrosis Initiation’ (NI), and ‘Full Necrosis’ (FN)), the SUV progressively decreased, with FN showing near-zero uptake (for mean values). Partial volume and PET noise effects were also evident in the quantitative data. The peak SUV was visibly lower for the 17 mm inserts compared to the 22 mm and 28 mm inserts for both tracers and the mean necrosis SUV values were always higher than 0 but lower than HT. All mean bias values for cross activity comparison for both tracers led to < 10% bias within limits of agreement (LoA) as shown in **Fig 5a**. Kernel Density Estimation (KDE) plots of voxel-level SUV values were created for the 28 mm inserts (**Fig 4**) clearly captured the designed uptake patterns for both tracers.

**Fig.5.**
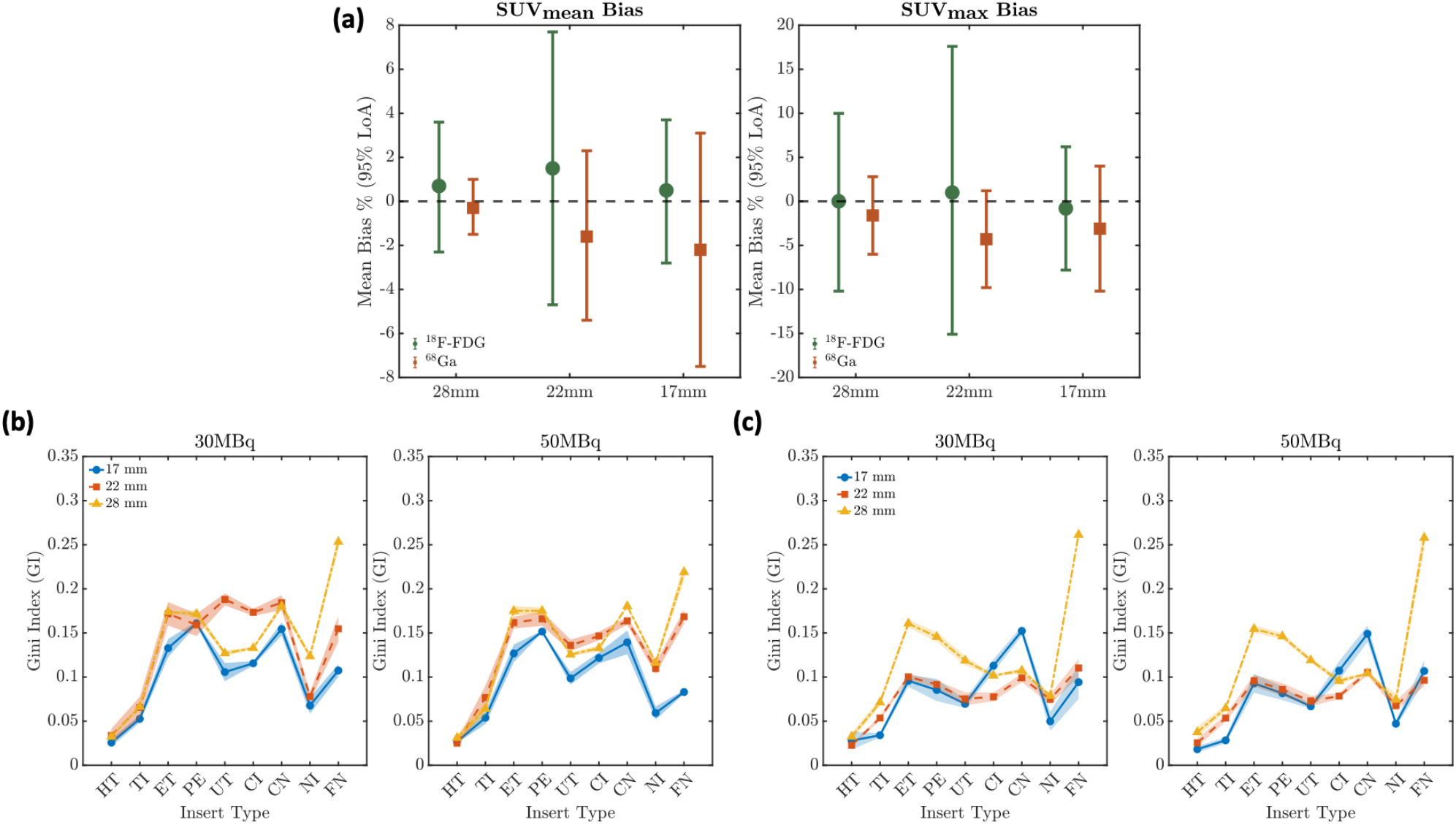
**(a)** Bland-Altman mean bias with 95% limits of agreement for 50 vs 30 MBq of extracted SUV values (Corresponding data in **Table A4**); Heterogeneity Gini index for **(b)** ^18^F-FDG & **(c)** ^68^Ga

### 3.2 PET Quantitative Metrics

Based on the phantom’s morphological design progression, the Gini index is expected to follow a characteristic non-linear trend: starting low for homogeneous targets (Healthy/Background), rising for targets with focal uptake inequalities (Incipient/Established), dipping for the spatially uniform target (Uniform Target), and rising again for targets exhibiting high-contrast asymmetry or necrosis (Crescent/Necrosis). The capability of the phantom to generate this quantifiable heterogeneity is illustrated in **Fig. 5b,c**, which plots the Gini Index as a function of insert design and VOI size. The GI values ranged between 0.05 – 0.3 with more uniform / low contrast heterogeneity inserts closer to 0.05.

To assess the design’s effectiveness, the target-to-background ratio (TBR_quant_) was evaluated and compared to the theoretical TBR_insert_ of ∼ 4. This analysis was performed on a 75% iso-value segmentation of the primary (hottest) target region, as defined in the methods. **Fig 6** plots the TBR_quant_ (mean and max) for the five inserts containing a distinct, hot primary region (ET, PE, UT, CI, CN). The other four inserts (HT, TI, NI, FN) were excluded from this specific analysis as they either lack a hot target (HT), have a very low-contrast target (TI), or contain necrosis (FN, NI), making them challenging to delineate reliably with a 75% threshold. The TBR_max_ values for the 28 mm inserts approached the theoretical TBR of 4. A qualitative comparison of the tracers showed that ^68^Ga (**Fig 6a, 6b**) consistently yielded a slightly lower TBR than ^18^F FDG (**Fig 6c, 6d**) for corresponding inserts and sizes. Furthermore, an analysis of TBR_max_ values relative to the 10% range of the theoretical TBR_insert_ (3.6 to 4.4) showed strong performance for the 28 mm inserts with all segmentations for both tracers fell within this range. This agreement degraded with size, as expected. On average the 28mm values fell with 10%, 22mm in 15-20% and 17mm in 30% bounds of TBR_insert._

**Fig.6.**
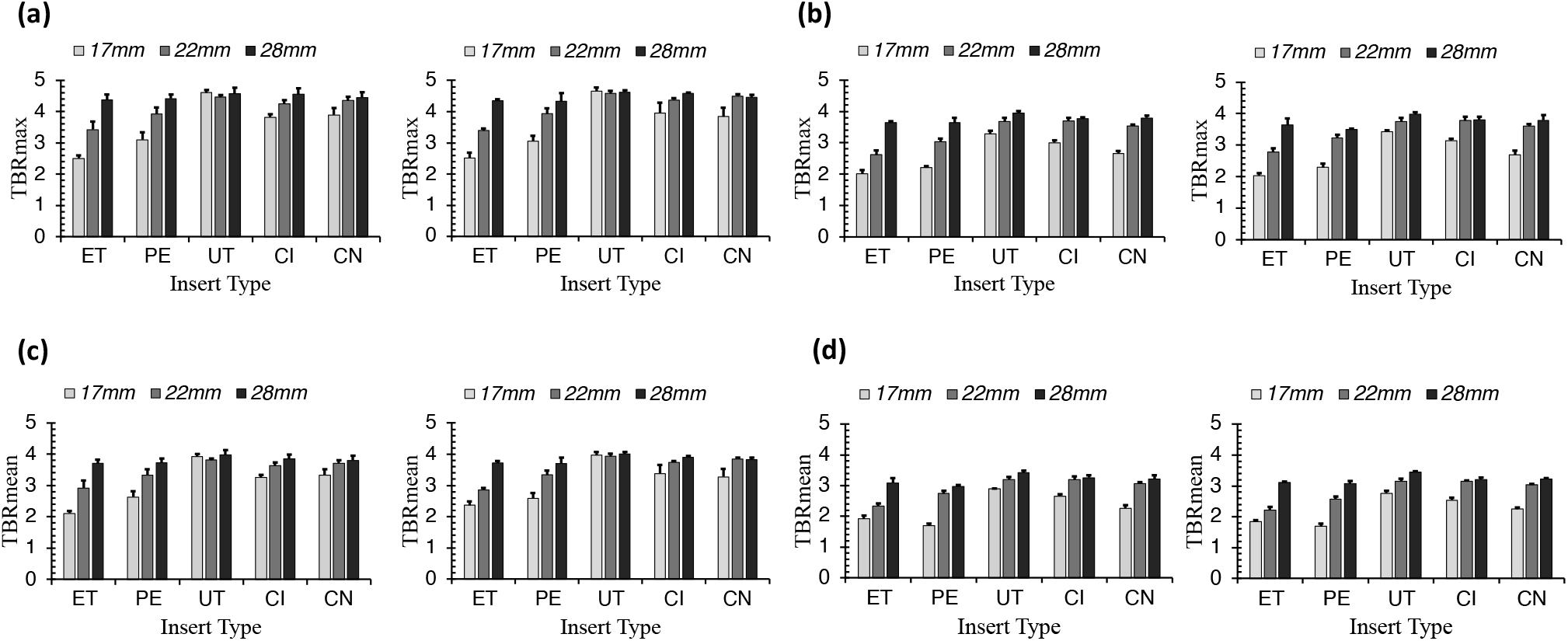
Target-to-Background Ratio (TBR_quant_) for the five hottest insert types, calculated from a 75% iso-value segmentation. Plots are shown for TBR_mean_ and TBR_max_ – (a) ^18^F-FDG; (b) ^68^Ga

## Discussion

This study successfully developed and validated a grid-inspired 3D-printed phantom inserts for simulating controlled spatial heterogeneous uptake patterns in PET imaging. The primary goal was to create a robust and reproducible tool that moves beyond simple hot-sphere phantoms with preparation simplicity. Hence, geometric simplicity (cubic outer shell, nested spherical voids) based on previous deign was chosen to ensure maximal quantitative interpretability and reproducibility as a prerequisite for future standardisation efforts. A crucial first step in validating the phantom’s quantitative integrity was to confirm the success of printing and uniformity of tracer mixing in the reference solution. The former was confirmed through 1.25mm CT air as shown in **Fig 2**. The analysis of the reference SUV_mean_ across the three phantom layers (**Table 2**) showed homogenous mixing for both tracers at both activity levels with a maximum variability of just 2.15%. Contrast to multi-compartment phantoms, proposed design functions with conventional acceptable standards of 10% **[10]** with a single tracer injection into the phantom body following findings of **[26]**.

The visual analysis of the PET images (**Fig 3a**) immediately confirmed the design’s success and its utility for studying the limits of spatial resolution. The intended heterogeneous patterns, such as crescent shapes and necrotic cores, were clearly discernible in the 28 mm inserts. These fine structures became less distinct in the 22 mm inserts and were visually indiscernible in the 17 mm inserts, collapsing into a more uniform-appearing hot spot. This is not a failure of the phantom but rather a key feature: it provides a clear, physical demonstration of partial volume effects (PVE) and the threshold of visual detectability for complex heterogeneity in small lesions. However, a critical finding is that despite this visual blurring, the quantitative SUV profiles (**Fig 3b-e**) for the all inserts still demonstrated the distinct trends of the designed progression from healthy (HT) to necrotic (FN). This illustrates a crucial clinical challenge: small lesions can appear visually homogeneous while possessing significant underlying quantitative heterogeneity **[27]**. The phantom’s ability to replicate this, is a significant advantage confirming that modulating the grid solidity is a highly effective method for controlling apparent tracer uptake. We acknowledge that clinical lesions often exhibit much higher contrast (typically TBR ∼ 8-10) than the TBR ∼ 4 used in this study. However, the primary goal was to build a reliable tool closer to the limits of detection and quantitative measurement, yet still practically achievable in terms of print resolution.

The voxel histograms in **Fig 4** reveal the distribution differences among the targets. While homogeneous targets (HT, UT) displayed symmetric peak (bell-shaped or flat peak), the heterogeneous inserts produced distinctly complex intensity profiles. Specifically, the multi-focal designs exhibited flattened, broad distributions indicating a mix of signal intensities, whereas necrotic models featured high-uptake peaks with extended tails stretching into the low-activity range. This contrast confirms the phantom’s ability to generate structural texture beyond simple averaging, with consistent voxel-level behaviour observed across both radiotracers. This suggests that the 3D-printed varying porosity successfully modulates the radiotracer distribution at a resolution detectable by the scanner. Furthermore, the pairwise statistical analysis (**Fig A1, A2**) validated that these simulated targets are not just visually different but are also quantitatively distinct except for 4 outliers.

We focused on validating the phantom’s quantitative integrity using standard, reproducible metrics such as SUV, GI, and TBR. The consistency of SUV trends between ^18^F-FDG and ^68^Ga, and across different activity concentrations is a crucial finding. This qualitative trend performance (**Fig 3b vs 3d**) demonstrates that the phantom’s relative contrast generation is robust, and independent of activity / tracer, making it a reliable tool for future standardization studies. Crucially, the mean SUV values for the necrotic regions were always higher than zero, reflecting the expected influence of printing resolution limitations and intrinsic PET noise/spill-in. The quantitative assessment of heterogeneity using the gini index revealed distinct trends correlating with the phantom’s morphological design. As illustrated in **Fig. 5**, the GI profiles generally exhibited a characteristic trend: starting low for the homogeneous insert, rising for the focal targets, dipping for the spatially uniform, and rising again for the asymmetric and necrotic targets. This non-linear behaviour confirms the metric’s sensitivity to structural complexity with the dip at the UT correctly distinguishing it from the surrounding heterogeneous grid structures. Full Necrosis frequently exhibited the highest Gini values (∼0.25). This spike is attributable to two compounding factors. First, the necrotic core represents a region of near-zero counts, creating a high-noise environment. Second, the inclusion of the outer boundary captures the high-contrast gradient between the cold target and the warm background. This creates maximal statistical inequality driving the Gini index. Certain outliers were also observed, in the lower-activity 30 MBq acquisition, the GI for the UT in the 22 mm inserts disrupted the expected dip. Finally, it is important to note that the observed Gini coefficients ranged from 0.05 to 0.30, well below the theoretical maximum of 1.0. This limited dynamic range is an expected consequence of the study design. As the phantom was engineered with TBR ∼ 4:1, the voxel intensity distribution is relatively tight leaded to tighter range for GI.

TBR_quant_ analysis (**Fig 6**) served to validate the design’s high-contrast performance against its theoretical TBR_insert_ of ∼ 4. The agreement for TBR_max_ in the 28 mm inserts confirmed the grid design is well-calibrated. The predictable degradation of TBR_quant_ (both mean and max) with decreasing insert size (22 mm and 17 mm) provided another clear, quantitative measure of PVE. Finally, the slight overestimation of TBR_max_ by ^18^F-FDG (especially in the UT insert) confirms a known bias in grid-based phantoms related to statistical noise **[15,19]**, which the more-blurred ^68^Ga signal was incidentally less susceptible to. Despite its demonstrated utility, this phantom study has inherent limitations that define the necessity of future work. This work focused on spatial heterogeneity and conventional PET metrics (SUV, GI, TBR) to validate the foundational ground truth. We did not perform a detailed radiomics analysis within the scope of this study. Although patterns like the crescent and necrotic shapes (CI, CN) were introduced, these models do not fully replicate the highly irregular or spiculated morphologies observed in real patient tumours. It is important to note that the exclusion of the lower-contrast inserts (TI, NI) from the segmentation validation, was an imposed limitation of the fixed 75% iso-surface threshold. This specific threshold, optimized for validating the highest contrast primary regions, does not reflect a failure of the phantom design to simulate these challenging patterns. Instead, it underscores the need for advanced adaptive segmentation algorithms that can reliably delineate low-contrast or near-zero uptake regions, confirming the phantom’s utility as a tool for developing and validating such methods. We acknowledge that full radiomic texture analysis is the next logical step. However, the primary aim of this manuscript is to validate the design feasibility and quantitative stability of the phantom. Establishing this ground truth is a prerequisite before evaluating complex radiomic feature stability. Crucially, the entire spatially heterogeneous structure for a given target size is produced as a single object from one design file, completely eliminating the multiple assembly steps or risk of leakage or cross-contamination associated with manually combining different regions of activity. Also, it is essential to note that this phantom is not intended for routine QC testing or system commissioning, which are adequately covered by current guidelines. Instead, it serves as a specialized additional tool for controlled heterogeneity simulation.

To conclude, the phantom provides a practical platform for radiomics studies where controlled, repeatable heterogeneity is essential. Known ground-truth patterns including cases where heterogeneity is quantitatively present but visually suppressed could be highly suited for evaluating feature robustness, sensitivity to partial-volume effects, segmentation uncertainty, and reconstruction biases. This tool could also support cross-site harmonization efforts by providing a standardized test object for feature stability benchmarking across centres, scanners, and protocols. It’s simple preparation workflow further enables large-scale, multi-institutional reproducibility studies.

## Conclusion

This study successfully developed and validated 3D-printed phantom inserts designed to simulate controlled spatial heterogeneity in PET imaging. The phantom provides a foundational ground-truth reference validated using conventional PET metrics.

- The tool demonstrates the quantitative presence of heterogeneity even when visually suppressed by PVE.
- Its reproducible nature across tracer supports potential for cross-site standardization efforts.

## Data Availability

All data produced in the present study are available upon reasonable request to the authors

## APPENDIX

Description for the nomenclature of these models shown in **main article (Fig 1.c)** is as below

- **Healthy (HT) :** Represents the baseline simulation of healthy tissue, characterized by a uniform 75% solidity
- **Target Incipient (TI) :** Simulates the earliest stage of lesion development, featuring a faint **primary region** with 50% infill. This initial region is typically smaller than the final intended target size – Multifocal Target
- **Established Target (ET) :** Simulates the first clearly detectable lesion. The *primary region*, maintaining the same size as in the IT stage, transitions to 0% infill, creating a distinct, hollow core - Multifocal Target
- **Progressive Expansion (PE) :** Simulates the active growth phase. A *secondary region* (outer ring) with 50% infill forms and expands around the hollow primary region established in the ET stage - Multifocal Target
- **Uniform Target (UT) :** Represents a single, homogeneous lesion that fills an entire intended target area (e.g., 17, 22, or 28 mm in diameter) – Uniform Target
- **Crescent Initiation (CI) :** Simulates a pattern forming within a Uniform Target. A new internal secondary region with 50% infill develops, causing the remaining original tissue to form a crescent-shaped primary region - Multifocal Target
- **Crescent Necrosis (CN) :** Simulates the progression of the crescent pattern. The internal **s**econdary region becomes fully necrotic, transitioning to 100% solidity, while being enclosed by the same crescent-shaped primary region – Necrosis Target
- **Necrosis Initiation (NI) :** Simulates the beginning of total lesion collapse in a target structure. The secondary region becomes fully necrotic (100% infill), while necrosis begins to consume the primary crescent region represented by a transition to 50% infill - Necrosis Target
- **Full Necrosis (FN) :** Represents the final end-stage. Both the primary and secondary regions are consumed, resulting in a single, homogeneous necrotic lesion emulation with 100% infill across the entire intended target size - Necrosis Target

**Table. A1.** Design Parameters Used for the Inserts.

| Healthy / Background | D<sub>UL</sub> in mm & Solidity %<br>(Primary Region) | D<sub>CC</sub> in mm & Solidity %<br>(Secondary Region) | D<sub>CR</sub> & Solidity %<br>(Secondary Region) |
| --- | --- | --- | --- |
| 0mm, 75% (HT) | 17mm, 0% (UT) | 11mm, 50% (TI) | 11mm, 50% (CI) |
|  | 17mm, 100% (FN) |  |  |
|  | 17mm, 50% (PE & NI) | 11mm, 0% (ET) | 11mm, 100% (CN) |
|  | 22 mm, 0% (UT) | 12mm, 50% (TI) | 12mm, 50% (CI) |
|  | 22mm, 100% (FN) |  |  |
|  | 22mm, 50% (PE & NI) | 12mm, 0% (ET) | 12mm, 100% (CN) |
|  | 28 mm, 0% (UT) | 17mm, 50% (TI) | 17mm, 50% (CI) |
|  | 28mm, 100% (FN) |  |  |
|  | 28mm, 50% (PE & NI) | 17mm, 50% (ET) | 17mm, 50% (CN) |

*NOTE: This preprint reports new research that is not certified by peer and should not be used to guide clinical practice.*

Recovery coefficient for the segmented data is calculated as below

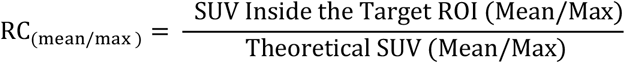

**Table. A2.**
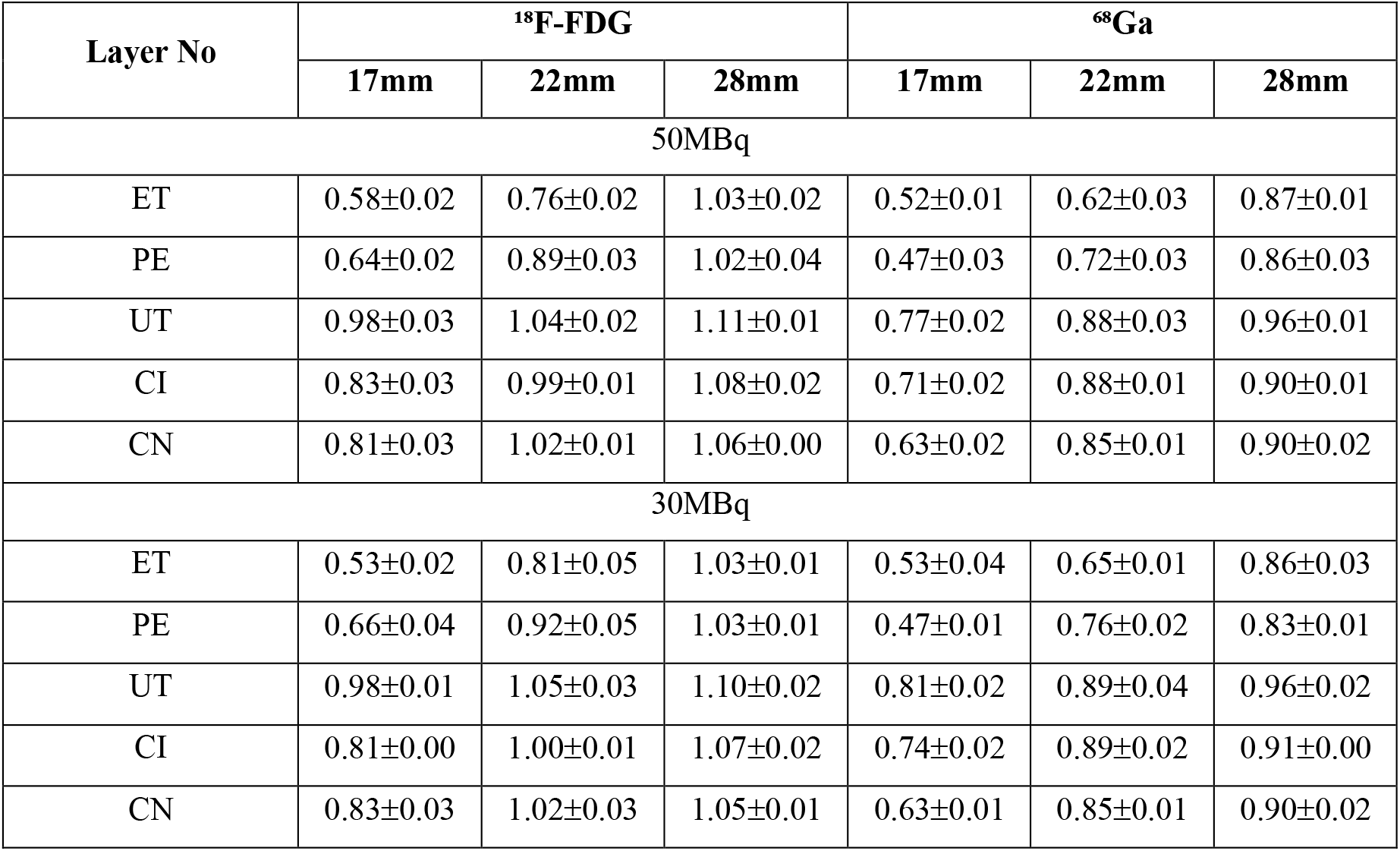
RC_mean_ calculated from segmented data for primary region.

**Table. A3.** RC_max_ calculated from segmented data for primary region.

| Layer No | <sup>18</sup>F-FDG |  |  | <sup>68</sup>Ga |  |  |
| --- | --- | --- | --- | --- | --- | --- |
|  | 17mm | 22mm | 28mm | 17mm | 22mm | 28mm |
| 50MBq |  |  |  |  |  |  |
| ET | 0.62±0.05 | 0.90±0.02 | 1.20±0.02 | 0.62±0.01 | 0.73±0.04 | 1.02±0.01 |
| PE | 0.75±0.02 | 1.03±0.04 | 1.20±0.05 | 0.56±0.04 | 0.85±0.03 | 1.02±0.05 |
| UT | 1.15±0.03 | 1.22±0.02 | 1.28±0.01 | 0.92±0.03 | 1.03±0.03 | 1.10±0.01 |
| CI | 0.98±0.04 | 1.16±0.01 | 1.26±0.03 | 0.84±0.02 | 1.04±0.02 | 1.05±0.01 |
| CN | 0.95±0.04 | 1.20±0.02 | 1.23±0.00 | 0.75±0.02 | 0.99±0.02 | 1.06±0.03 |
| 30MBq |  |  |  |  |  |  |
| ET | 0.62±0.03 | 0.94±0.05 | 1.21±0.02 | 0.64±0.04 | 0.77±0.02 | 1.01±0.04 |
| PE | 0.77±0.05 | 1.08±0.06 | 1.22±0.01 | 0.57±0.01 | 0.90±0.02 | 0.98±0.01 |
| UT | 1.15±0.01 | 1.23±0.04 | 1.27±0.02 | 0.95±0.04 | 1.04±0.05 | 1.11±0.04 |
| CI | 0.95±0.00 | 1.17±0.01 | 1.26±0.04 | 0.88±0.00 | 1.05±0.03 | 1.06±0.01 |
| CN | 0.97±0.04 | 1.20±0.05 | 1.23±0.02 | 0.75±0.02 | 1.00±0.01 | 1.06±0.02 |

*NOTE: This preprint reports new research that is not certified by peer and should not be used to guide clinical practice.*

**Table. A4.**
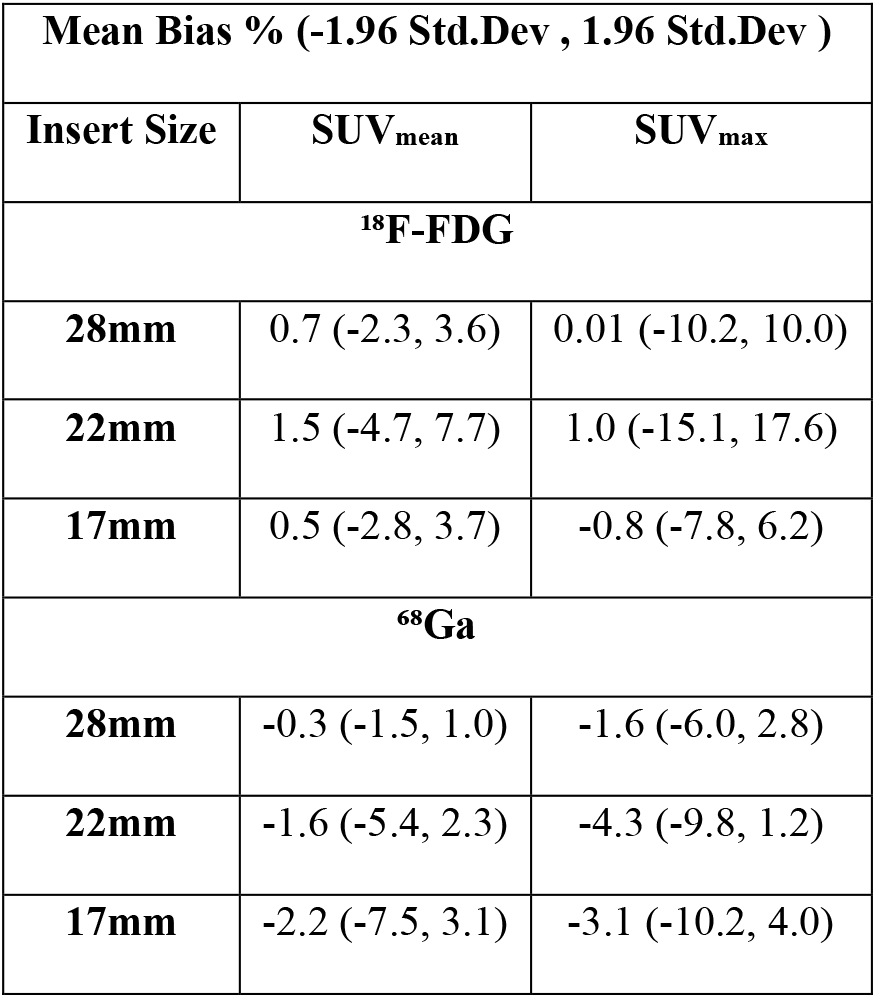
Bland-Altman Mean Bian with Limits of Agreement in % for 50 vs 30 MBq.

**Fig.A1.**
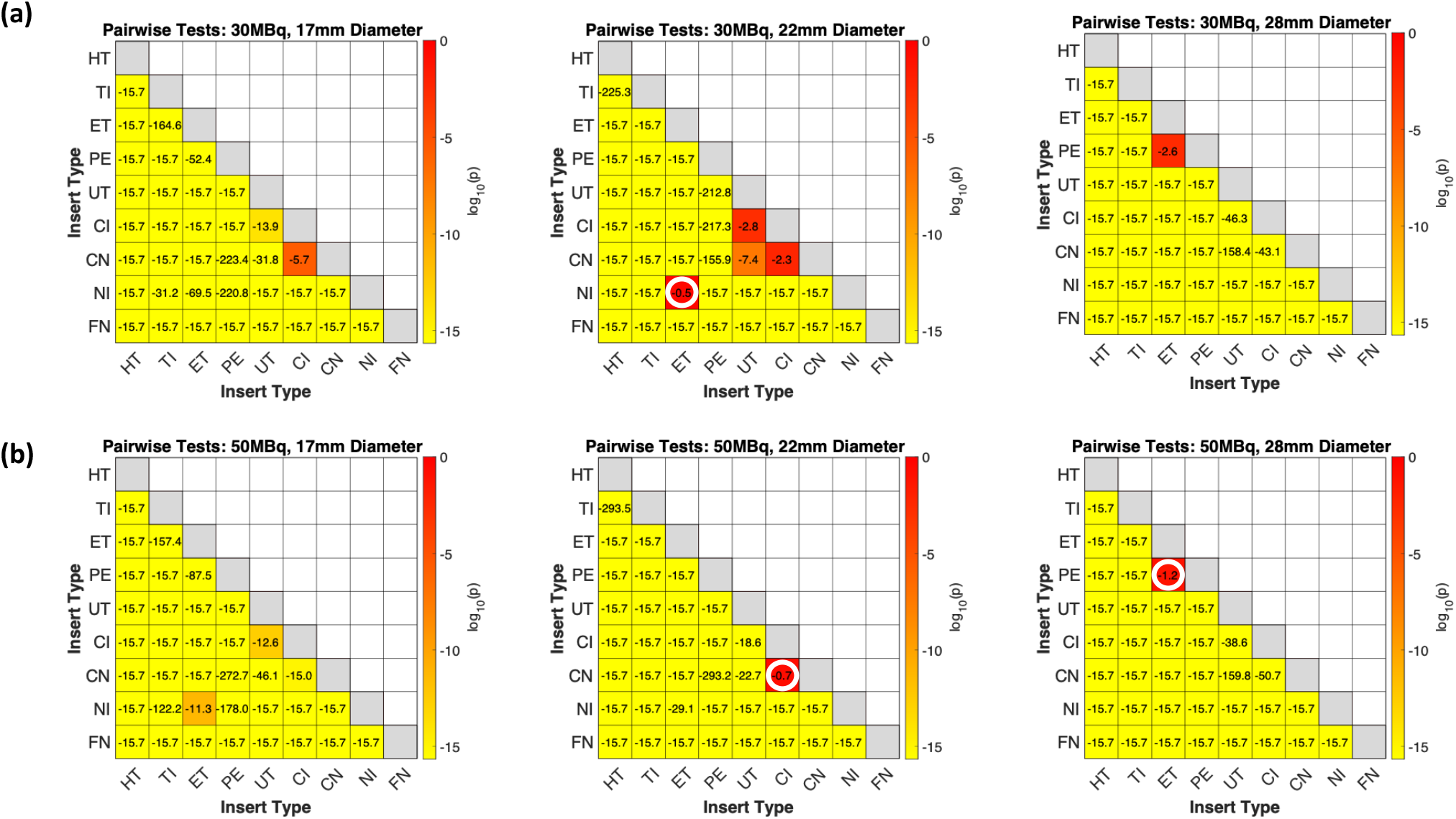
Wilcoxon Rank Sum test for the VOI histogram data of 28mm inserts for [^18^F]FDG **(a)** 30Mbq**; (b)** 50MBq – p value plotted in log scale : **≤ -1.3** which is log(0.05) - (Values higher than -1.3 indicates failures i.e. the two targets are statistically similar to each other)

**Fig.A2.**
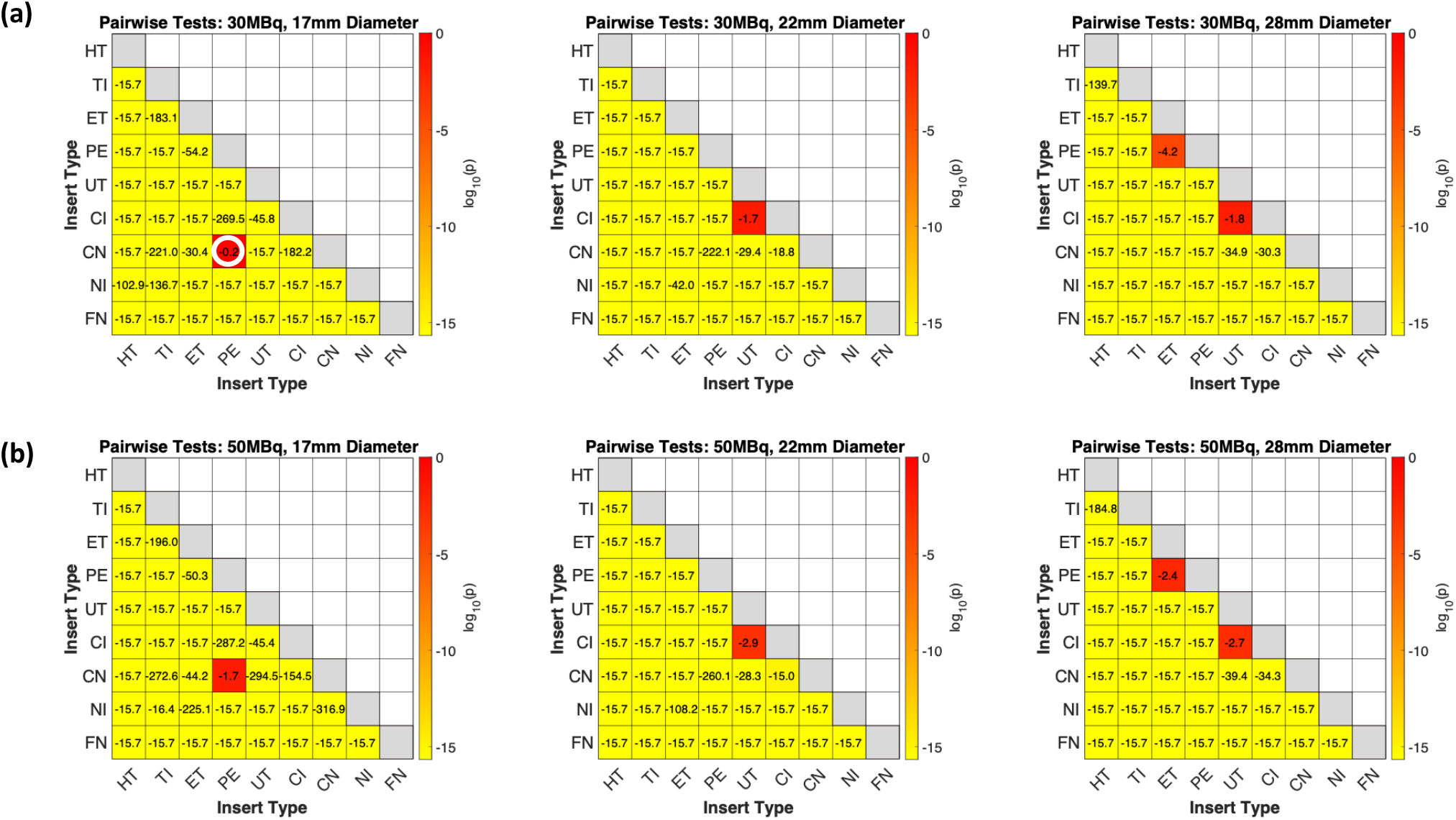
Wilcoxon Rank Sum test for the VOI histogram data of 28mm inserts for [^68^Ga] **(a)** 30Mbq**; (b)** 50MBq – p value plotted in log scale : **≤ -1.3** which is log(0.05) - (Values higher than -1.3 indicates failures i.e. the two targets are similar to each other)

